# Persistent Autoimmune Activation and Proinflammatory State in Post-COVID Syndrome

**DOI:** 10.1101/2021.11.17.21266457

**Authors:** Yeny Acosta-Ampudia, Diana M Monsalve, Manuel Rojas, Yhojan Rodríguez, Elizabeth Zapata, Carolina Ramírez-Santana, Juan-Manuel Anaya

## Abstract

**Background:** The immunopathological pathways enabling post-COVID syndrome (PCS) development are not entirely known. We underwent a longitudinal analysis of patients with COVID-19 who developed PCS aiming to evaluate the autoimmune and immunological status associated with this condition.

**Methods:** Thirty-three patients were included for longitudinal clinical and autoantibody analyses of whom 12 patients were assessed for cytokines and lymphocyte populations. Patients were followed during 7-11 months after acute COVID-19. Autoimmune profile and immunological status were evaluated mainly by enzyme-linked-immunosorbent assays and flow cytometry.

**Results:** Latent autoimmunity and overt autoimmunity persisted over time. A proinflammatory state was observed in patients with PCS characterized by upregulated IFN-α, TNF-α, G-CSF, IL-17A, IL-6, IL-1β, and IL-13, whereas IP-10 was decreased. In addition, PCS was characterized by increased levels of Th9, CD8+ effector T cells, naive B cells, and CD4+ effector memory T cells. Total levels of IgG S1-SARS-CoV-2 antibodies remained elevated over time.

**Discussion:** The clinical manifestations of PCS are associated with the persistence of a proinflammatory, and effector phenotype induced by SARS-CoV-2 infection. This long-term persistent immune activation may contribute to the development of latent and overt autoimmunity. Results suggest the need to evaluate the role of immunomodulation in the treatment of PCS.

## Introduction

During acute infection of severe acute respiratory syndrome coronavirus 2 (SARS-CoV-2), responsible for the coronavirus disease 2019 (COVID-19), clinical manifestations vary from mild forms to critical and more severe cases [1]. Symptoms include dry cough, fatigue, anosmia, and fever. However, some patients may worsen requiring intensive care unit (ICU) admission, mechanical ventilation and vasopressor support [1].

Although most of the COVID-19 patients recover entirely, without sequelae, some patients may keep experiencing symptoms after four weeks of acute disease and others may even develop new symptoms [2]. This clinical spectrum is called post-COVID syndrome (PCS) [2]. One-third of patients with PCS present with at least one musculoskeletal, respiratory, gastrointestinal and neurological symptoms [2]. The causes of PCS are under study. Viral persistence [3], endocrine dysregulation [4], endotheliopathy [5], autoimmune response [6], and a persistent inflammatory state [7], have been advocated. However, the precise mechanisms associated with its appearance, and the influence of biological alterations on clinical phenotypes remain to be elucidated. Herein we present a longitudinal analysis of a case series of patients with COVID-19 who developed PCS aiming to evaluate the autoimmune and immunological pathways associated with this condition, and the likely targets for novel treatments.

## Methods

### Study design

COVID-19 patients who participated in a previous study [8], were contacted by telephone and reassessed for possible post-COVID symptoms. Those who were suspected to present PCS were invited to attend the post-COVID unit at the Clínica del Occidente, in Bogota, Colombia, for clinical and immunological evaluation. A final sample size of 33 patients was included for longitudinal autoantibody analyses and 12 were assessed for cytokines, lymphocyte populations and anti-SARS-CoV-2 antibodies (Figure 1A). Serological data of 100 patients, previously reported, served as historical control group for autoimmune comparisons [9], and a group of 8 pre-pandemic healthy individuals was the control group for the immunological assessment [8].

**Figure 1.**
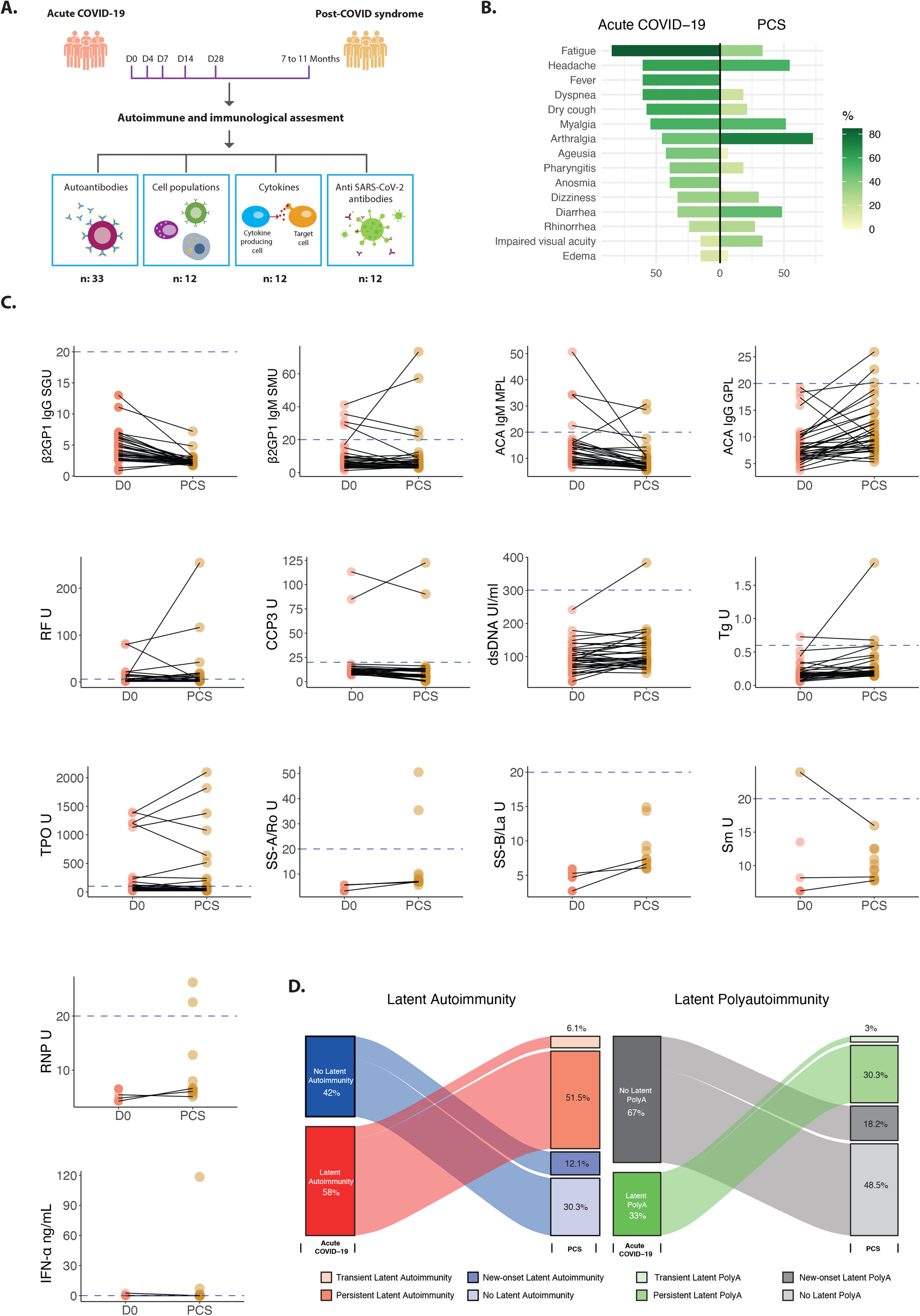
Autoimmune assessment of post-COVID syndrome. **A**. Study design. Patients assessed for autoantibodies were followed 7-11 months post infection (n: 33). Patients evaluated for cytokines, lymphocytes, IgG, and IgA S1-SARS-CoV-2 antibodies were followed 7-9 months post infection (n: 12). **B**. Mirrored bar plot for symptoms on acute COVID-19 and post-COVID syndrome (n:33). **C**. Paired dot plot for concentration of autoantibodies. Dashed blue line represents cutoff values for positivity of each autoantibody. **D**. Alluvial diagrams for latent autoimmunity and overt autoimmunity. Following are the abbreviations for autoantibodies: ACAs: Anti-cardiolipin antibodies; β2GPI: β2 glycoprotein-1; CCP3: Cyclic citrullinated peptide third-generation; dsDNA: Double-stranded DNA; IFN-α: Interferon-α; RF: Rheumatoid factor; RNP: Ribonucleoprotein; Sm: Smith; Tg: Thyroglobulin. COVID-19: Coronavirus disease 2019; Ig: Immunoglobulin; PolyA: Polyautoimmunity; PCS: Post-COVID syndrome.

### Patient monitoring and clinical evaluation

Patients were systematically evaluated for acute COVID-19 and post-COVID clinical manifestations, as previously described [2]. Out of 33 patients with PCS evaluated for autoantibodies, five were vaccinated during the PCS. This study was done in compliance with Act 008430/1993 of the Ministry of Health of the Republic of Colombia, which classified it as minimal-risk research. All the patients were asked for their consent and were informed about the Colombian data protection law (1581 of 2012). The institutional review board of the CES University approved the study design.

### Autoantibodies

Detection of IgM rheumatoid factor (RF), IgG anti-cyclic citrullinated peptide third generation (CCP3), IgM and IgG anti-cardiolipin antibodies (ACAs), IgM and IgG anti-β2 glycoprotein-1 (β2GP1) antibodies, IgG anti-double-stranded DNA (dsDNA) antibodies, IgG anti-thyroglobulin (Tg) antibodies and anti-thyroid peroxidase (TPO) antibodies were all quantified by enzyme-linked-immunosorbent assay (ELISA). In addition, antinuclear antibodies (ANAs) were evaluated by using an indirect immunofluorescence assay. Positive results were considered from dilution 1/80. In case of ANAs positivity, anti-SSA/Ro, anti-SSB/La, anti-ribonucleoprotein (RNP) and anti-smith (Sm) antibodies were further evaluated by a commercial ELISA. All the assay kits were from Inova Diagnostics, Inc (San Diego, CA, USA) as previously reported [9]. Assessment of anti-IFN-α antibodies was done by ELISA (Thermo Fisher Scientific, Waltham, MA) following manufacturer’s specifications.

### Cytokine assay and lymphocytes immunophenotype

Serum concentration of 20 cytokines (IL-1β, IL-2, IL-4, IL-5, IL-6, IL-7, IL-8, IL-9, IL-10, IL-12p70, IL-13, IL-17A, TNF-α, G-CSF, GM-CSF, RANTES, MCP-1, IP-10, IFN-γ, IFN-α) was assessed by Cytometric Bead Array (CBA, Becton Dickinson Biosciences, San Diego, CA, USA). The test was done according to the manufacturer’s protocols. Concentration of the cytokines was calculated using the FCAP Array™ Software (BD Bioscience) as reported elsewhere [10].

Thirty cell subsets (Supplementary Appendix) were also assessed. A minimum of 100,000 lymphocytes per sample were acquired on a FACSCanto II™ flow cytometer (BD Biosciences™) and data was analyzed with FlowJo software version 9 (BD Biosciences™) as reported elsewhere [8].

### Anti-SARS-CoV-2 Antibodies

The Euroimmun anti-SARS-CoV-2 ELISA (Euroimmun, Luebeck, Germany) was used for serological detection of human IgG and IgA antibodies against the SARS-CoV-2 S1 structural protein, in accordance with the manufacturer’s instructions, as previously described [2]. The ratio interpretation was <0.8 = negative, ≥0.8 to <1.1 = borderline, ≥ 1.1 = positive. Antibody positivity was performed using a 1:100 dilution.

### Statistical analysis

Univariate descriptive statistics were performed. Categorical variables were analyzed using frequencies, and quantitative continuous variables were expressed in the median and interquartile range (IQR). Fisher’s exact tests were used based on the results. Cytokine concentrations were analyzed after log transformation; all other parameters were analyzed without any additional data transformation.

Initially, generalized linear models were used to evaluate longitudinal changes in IgA and IgG SARS-CoV-2 antibodies ratios, cytokines, and lymphocyte populations, as previously described [8]. In addition, linear regression models were fitted to estimate the differences in cytokines and lymphocyte populations between PCS and pre-pandemic controls. For these models, *post hoc* comparison of means was based on both adjusted Bonferroni p-values and Fisher’s protected least significant differences procedure using t statistics based on Satterwhaite’s approximation.

Next, we evaluated whether levels and positivity of autoantibodies changed from acute COVID-19 to PCS. The McNemar test with continuity correction and paired T-test were performed for positivity and autoantibodies levels, respectively. The significance level of the study was set to 0.05. Statistical analyses were done using R software version 4.0.2.

## Results

### Autoimmune assessment

The main clinical characteristics of the 33 patients are shown in Figure 1B. Most of them were male (19/33, 57.6%) with a median age of 55 years (IQR: 50 to 63). The median post-COVID time was 266 (IQR: 253 to 288) days.

Concentration of ACA IgG (P= 0.0013), dsDNA (P= 0.0108), and Tg (P= 0.03019) autoantibodies increase from day 0 to PCS (Figure 1C). Whereas, ACA IgM (P= 0.0026), and β2GP1 IgG (P <.0001) autoantibodies declined (Figure 1C). Other autoantibodies concentration did not fluctuate during the follow-up (Paired T-test, P >.0500). Despite the variation in concentration, autoantibodies positivity did not change from acute disease (D0) to PCS (McNemar test, P >0.0500), except for ANAs which showed a slight increase in its frequency at 1/80 dilution (McNemar test, P= 0.0455).

As compared with pre-pandemic controls, frequency of β2GP1 IgM autoantibodies was higher in PCS (Fisher’s exact test, P=0.0135). The remaining autoantibodies did not differ from healthy subjects at baseline (D0) or at the time of PCS (Fisher’s exact test, P >.0500).

During the acute phase of illness there were 19/33 (57.6%) patients presenting with at least one autoantibody (i.e., latent autoimmunity), of whom 11/33 (33.3%) patients presented with two or more autoantibodies (i.e., latent polyautoimmunity), while during the PCS there were 21/33 (63.6%) patients presenting with at least one autoantibody (Fisher’s exact test, P=0.0006; McNemar test, P= 0.6831), of whom 16/33 (48.5%) presented with two or more autoantibodies (Fisher’s exact test, P=0.0007; McNemar test, P= 0.1306) (Figure 1D). These results indicate that there was not only a persistence of latent autoimmunity during the PCS but also an increased prevalence of both latent autoimmunity and latent polyautoimmunity (Figure 1D).

At the acute COVID-19 5/33 (15.2%) patients disclosed non-autoimmune hypothyroidism, and 3/33 (9.1%) had autoimmune thyroid disease (AITD). During the PCS, there was a patient who developed AITD, and other in whom anti-TPO antibodies became negative.

### Immunological assessment

The main clinical characteristics of the 12 patients are shown in Figure 2A. Half were male (6/12, 50%) with a median age of 50.5 years (IQR: 49.75 to 55.5). The median post-COVID time was 259 (IQR: 235.5 to 271.5) days. Although during the acute COVID-19, patients presented a progressive increase of IgA (from day 0 to day 28, P=0.0257) and IgG (from day 0 to day 28, P<.0001) anti-SARS-CoV-2 S1 antibodies, a reduction of 22.3% and 33.6% was observed at the PCS evaluation, respectively. However, IgA antibodies returned to similar levels from baseline (P= 1.0000), and IgG antibodies remained high (P <.0001) (Figure 2B).

**Figure 2.**
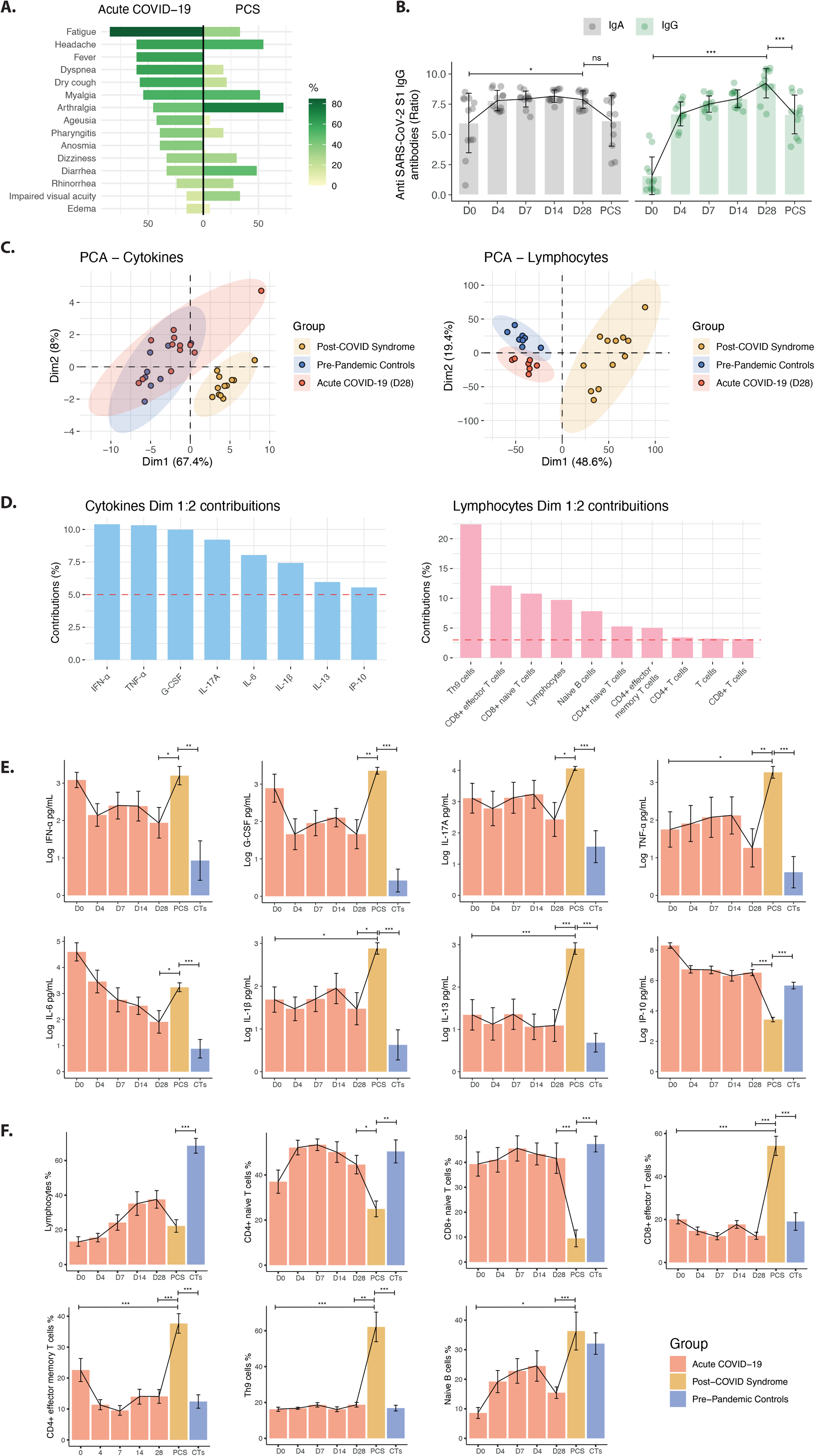
Immunological assessment of post-COVID syndrome. **A**. Mirrored bar plot for symptoms of acute COVID-19 and post-COVID syndrome (n:12). **B**. Dynamics of the SARS-CoV-2 antibody response and the change in anti-SARS-CoV-2 IgA and IgG ratios (OD sample/OD calibrator) were plotted against days of follow-up. **C**. Principal component analysis of 20 cytokines and 30 cell subsets that were analyzed in pre-pandemic controls (n: 8), acute COVID-19 (n:12), and PCS (n: 12). **D**. Contribution of cytokines and lymphocytes on dimensions 1 and 2 from principal component analyses. Thresholds for cytokines 5% (1/20 cytokines = 5%), and lymphocytes (1/30 populations = 3.3%). **E**. Longitudinal bar plots for selected cytokines in pre-pandemic controls (n: 8), acute COVID-19 (n:12), and PCS (n: 12). Longitudinal analyses were done by generalized linear models with *post hoc* comparisons adjusted by Bonferroni correction. Comparisons between pre-pandemic controls and PCS were analyzed by means of linear regression with *post hoc* comparison. **F**. Longitudinal bar plots for selected lymphocyte populations in pre-pandemic controls (n: 8), acute COVID-19 (n:12), and PCS (n: 12). Longitudinal analyses were done by generalized linear models with *post hoc* comparisons adjusted by Bonferroni correction. Comparisons between pre-pandemic controls and PCS were analyzed by means of linear regression with *post hoc* comparison. *** P < 0.0001, ** P < .0010, * P <.0500, ns: Not significant. PCS: Post-COVID syndrome; COVID-19: Coronavirus disease 2019; CTs: Pre-pandemic controls; D: Days; PCA: Principal components analysis; SARS-CoV-2: Severe acute respiratory syndrome coronavirus 2; G-CSF: Granulocyte colony-stimulating factor; IFN: Interferon; IL: Interleukin; Ig: Immunoglobulin; IP-10: interferon-γ induced protein 10; TNF-α: Tumor necrosis factor-alpha.

### Cytokine and lymphocyte profiles in PCS

Principal component analysis (PCA) of a panel of cytokine and lymphocyte populations contributing to PCS pathophysiology showed that PCS patients were segregated from the other two groups of patients (i.e., day 28 and pre-pandemic controls) (Figure 2C). Then, the most critical factors for such distinction were assessed. It was found that IFN-α, TNF-α, G-CSF, IL-17A, IL-6, IL-1β, IL-13, and IP-10 were the most contributing cytokines (i.e., average contribution cutoff >5%) (Figure 2D). The most contributing lymphocyte immunophenotypes were Th9, CD8+ effector T cells, CD8+ naive T cells, total lymphocytes, naive B cells, CD4+ naive T cells, CD4+ effector memory T cells, CD4+ and CD8+T cells, and total T cells (average contribution cutoff >3.33%) (Figure 2D).

Longitudinal evaluation of selected biomarkers disclosed an increase of most of the cytokines during PCS compared with levels measured at day 28 and with pre-pandemic controls, whereas IP-10 decreased (Figure 2E). TNF-α (P= 0.0118), IL-1β (P= 0.0166), and IL-13 (P <.0001) levels were higher during PCS than at day 0 (Figure 2E).

Regarding lymphocytes, Th9, CD8+ effector T cells, naive B cells, and CD4+ effector memory T cells were increased in PCS compared with levels measured at day 28 and with pre-pandemic controls (Figure 2F). On the other hand, lower levels of CD8+ and CD4+ naive T cells, and total lymphocytes, were observed (Figure 2F). However, these variations did not reach statistical significance in both groups.

## Discussion

This study shows the development of autoimmunity during PCS (Figure 1D) together with a persistent proinflammatory state and a dysregulated cellular immune response.

Levels of IgG anti-SARS-CoV-2 S1 antibodies remained high since the acute phase of illness to the PCS, while IgA antibodies tend to return to the basal state. Long-term antibody responses to SARS-CoV-2 and a high inter-individual variability has been reported [2]. Other longitudinal studies have shown small or no change in neutralizing antibody titers at 5 months post-infection [11,12]. Chen et al.

[13] showed that 92.5% of patients had detectable neutralizing antibodies approximately 7 months after infection, despite a decrease in antibody titers. This is in line with our study, in which IgG and IgA antibodies against S1 SARS-CoV-2 decreased slightly 7 to 9 months of evaluation. Similarly, a study conducted by Ivanov et al. [14] in which asymptomatic or mild COVID-19 patients were evaluated, showed that levels of IgA antibodies persisted high for more than 6 months after recovery from COVID-19.

Latent polyautoimmunity is associated with deleterious outcomes in patients with acute COVID-19 [9], and persistent of autoantibodies could influence clinical phenotypes in PCS, and the development of overt autoimmunity [15]. Our results demonstrate that latent polyautoimmunity observed during the acute phase of disease persists during PCS. In other words, latent polyautoimmunity in COVID-19 is not transient. Persistent ACA IgG positivity has been reported in a patient developing PCS [16], as well as an increase in the positivity of ANAs antibodies during PCS [17]. Nevertheless, since no information of autoantibodies before SARS-CoV-2 infection exist, a causal effect of virus infection in the development of autoimmunity during the acute phase of disease is precluded.

Cañas et al. [6] proposed that the development of autoimmune conditions subsequent to COVID-19 infection could be associated with transient immunosuppression of innate and acquired immunity leading to a loss of self-tolerance, and with a form of inappropriate immune reconstitution in genetically susceptible individuals. Our results show that patients with PCS have high levels of naive B cells, which are known to be a source of autoantibodies [18,19].

We observed an increase of proinflammatory cytokines in PCS (i.e., IFN-α, TNF-α, G-CSF, IL17A, IL-6, IL1-β and IL-13) confirming previous observations [20,21]. In addition, a significant decrease in IP-10 was noticed. This cytokine is a biomarker associated with severity and risk of death in COVID-19 patients. IP-10 is implicated in the acute phase of COVID-19 as promoting viral clearance and as an effector of immune-mediated acute lung injury [22]. Our results suggest that IP-10 plays a critical role in the acute phase of illness but not in PCS.

Studies based on the general population have shown associations between high concentrations of circulating inflammatory markers, such as C-reactive protein (CRP), IL-6, and TNF-α with depressive symptoms [23,24]. Specifically, IL-6 has been associated with central and peripheral nervous system complications [25,26]. In PCS, persistent IL-6 dysregulation may contribute to fatigue, sleeping difficulties, depression, and anxiety suggesting that a maintained inflammation is associated with these symptoms. IL-6 and other cytokines can signal the brain via volume diffusion in circumventricular zones, afferent nerves, and by active blood-brain-barrier transport [27].

IL-6 influences memory processes such as long-term potentiation, depression and promote and regulate sleep-related processes in PCS [26]. On the other hand, a systematic review on major depressive disorder biomarkers in PCS, showed increased levels of IL-6, IL-1β, TNF-α, IFN-γ, IL-10, IL-2, CRP, MCP-1, serum amyloid A and metabolites of the kynurenine pathway [28]. The identification of these biomarkers, associated with chronic inflammation, could help to understand the pathophysiology of depression in PCS. Moreover, IL-6, could drive autoinflammatory reactions and autoimmunity, via pre-existing natural B cell clones [29].

Regarding cellular immune response, our results reveal that 7 to 9 months after SARS-CoV-2 infection, most of the components of cellular immunity do not return to normal baseline in patients with PCS. An increase in CD4+ effector memory T cells, CD8+ effector T cells, Th9, and naive B cells was observed.

Studies in acute COVID-19 have shown a functional depletion and decrease in CD4+ and CD8+ T lymphocytes, and natural killer cells numbers [30]. The apoptosis induced by SARS-CoV-2 in lymphocytes expressing the angiotensin-converting enzyme 2 receptor and the cytokine storm explain this phenomenon [30]. In our study, we found that lymphopenia persists in PCS, affecting total lymphocytes, and CD8+ and CD4+ naive T cells. In addition to the apoptosis process, this lymphopenia can be caused by a change of the CD4+ and CD8+ T cell effector phenotype. Townsend et al. [31], demonstrated an expansion of effector CD8+ T cells and activated CD4+ and CD8+ T cells, with a reduction in naive CD4+ and CD8+ T cells at 101 days post-infection. Varghese et al. [32] showed that lymphopenia persisted in 14% of patients 102 days post-infection. A significant increase in Th9 cells compared to pre-pandemic controls and COVID-19 acute phase (i.e., D28) was observed in our study. Previous studies described that Th9 responses mediated by IL-9 production are involved in tissue inflammation and immune-mediated diseases such as autoimmunity and asthma [33]. Orologas-Stavrou et al. [34], showed a high Th9/Th17 ratio associated with persistence of a generalized inflammatory reaction (by Th17 cells), particularly in lungs (by Th9 cells) two months after infection.

Our study has several strengths. The longitudinal follow-up allows studying the causality of SARS-CoV-2 infection on the clinical and immunological profiles evaluated several months after infection. This allowed us to compare clinical and immunological phenotypes during PCS. In addition, pre-pandemic controls guarantee the lack of influence of unrecognized/asymptomatic infections on the autoimmune and immunological profiles. None of the included patients received immunomodulatory treatments that may had influenced the immune response during PCS.

Study limitations are acknowledged. A broader panel of B and T cells, as well as their functional analyses was not possible. Genetic analysis was not done. Another potential shortcoming of the present study is that the observed results might be due to chance alone or to the moderate sample size. However, this is unlikely because of the highly significant results observed after adjustments and corrections as well as their consistent direction and magnitude within the different analyses.

## Conclusions

The clinical manifestations in PCS could be associated with the persistence of a proinflammatory and effector phenotype induced by SARS-CoV-2 infection. Results advocate for therapeutic targets for management of PCS, including TNF or IL-6 blockade, and immunomodulatory drugs. Additionally, this long-term persistent immune activation could contribute to reactivity and the development of latent and overt autoimmunity.

## Data Availability

All data produced in the present study are available upon reasonable request to the authors

## Abbreviations

ACAs: Anti-cardiolipin antibodies.
AITD: Autoimmune thyroid disease.
ANAs: Anti-nuclear antibodies.
β2GP1: β2 glycoprotein-1.
CCP3: Citrullinated peptide third generation.
COVID-19: Coronavirus disease 2019.
CRP: C reactive protein.
dsDNA: Double-stranded DNA.
ELISA: Enzyme-linked-immunosorbent assay.
ICU: Intensive care unit.
IQR: Interquartile range.
PCA: Principal component analysis.
PCS: Post-COVID syndrome.
RF: Rheumatoid Factor.
RNP: Ribonucleoprotein.
SARS-CoV-2: Severe acute respiratory syndrome coronavirus 2.
Sm: Smith.
Tg: Thyroglobulin.
TPO: Thyroid peroxidase.

## Declaration of competing interest

None.

## Funding

This work was supported by the Universidad del Rosario grant numbers IV-FBG001, and ABN-011.

## Role of the Funder/Sponsor

The funders had no role in the design and conduct of the study, collection, management, analysis, and interpretation of the data; preparation, review, or approval of the manuscript; and decision to submit the manuscript for publication.

## Acknowledgments

The authors would like to thank all the members of the CREA for their contributions and fruitful discussions during the preparation of the manuscript.

## Notes

### Competing Interest Statement

The authors have declared no competing interest.

### Author Declarations

The institutional review board of the CES University approved the study design.

## References

1. Dhama K, Khan S, Tiwari R, et al. Coronavirus Disease 2019–COVID-19. Clin Microbiol Rev. 2020; 33(4).

2. Anaya J-M, Rojas M, Salinas ML, et al. Post-COVID syndrome. A case series and comprehensive review. Autoimmun Rev. 2021; 20(11):102947.

3. Jacobs JJL. Persistent SARS-2 infections contribute to long COVID-19. Med Hypotheses. 2021/02/16. Published by Elsevier Ltd.; 2021; 149:110538.

4. Mongioì LM, Barbagallo F, Condorelli RA, et al. Possible long-term endocrine-metabolic complications in COVID-19: lesson from the SARS model. Endocrine. 2020/06/02. Springer US; 2020; 68(3):467–470.

5. Fogarty H, Townsend L, Morrin H, et al. Persistent endotheliopathy in the pathogenesis of long COVID syndrome. J Thromb Haemost. 2021; 19(10):2546–2553.

6. Cañas CA. The triggering of post-COVID-19 autoimmunity phenomena could be associated with both transient immunosuppression and an inappropriate form of immune reconstitution in susceptible individuals. Med Hypotheses. 2020/10/14. Elsevier Ltd.; 2020; 145:110345.

7. Tserel L, Jõgi P, Naaber P, et al. Long-Term Elevated Inflammatory Protein Levels in Asymptomatic SARS-CoV-2 Infected Individuals. Front Immunol. 2021; 12:709759.

8. Acosta-Ampudia Y, Monsalve DMM, Rojas M, et al. COVID-19 convalescent plasma composition and immunological effects in severe patients. J Autoimmun. 2021; 118:102598.

9. Anaya J-M, Monsalve DM, Rojas M, et al. Latent rheumatic, thyroid and phospholipid autoimmunity in hospitalized patients with COVID-19. J Transl Autoimmun. 2021; 4:100091.

10. Pacheco Y, Barahona-Correa J, Monsalve DM, et al. Cytokine and autoantibody clusters interaction in systemic lupus erythematosus. J Transl Med. 2017; 15(1):239.

11. Wajnberg A, Amanat F, Firpo A, et al. Robust neutralizing antibodies to SARS-CoV-2 infection persist for months. Science. 2020/10/28. American Association for the Advancement of Science; 2020; 370(6521):1227–1230.

12. Iyer AS, Jones FK, Nodoushani A, et al. Persistence and decay of human antibody responses to the receptor binding domain of SARS-CoV-2 spike protein in COVID-19 patients. Sci Immunol. 2020; 5(52).

13. Chen J, Liu X, Zhang X, et al. Decline in neutralising antibody responses, but sustained T-cell immunity, in COVID-19 patients at 7 months post-infection. Clin Transl Immunol. 2021; 10(7):e1319.

14. Ivanov A, Semenova E. Long-term monitoring of the development and extinction of IgA and IgG responses to SARS-CoV-2 infection. J Med Virol. 2021; 93(10):5953–5960.

15. Knight JS, Caricchio R, Casanova JL, et al. The intersection of COVID-19 and autoimmunity. J Clin Invest [Internet]. The American Society for Clinical Investigation; 2021;. Available from: https://doi.org/10.1172/JCI154886

16. Bertin D, Kaphan E, Weber S, et al. Persistent IgG anticardiolipin autoantibodies are associated with post-COVID syndrome. Int. J. Infect. Dis. IJID Off. Publ. Int. Soc. Infect. Dis. 2021. p. 23–25.

17. Seeßle J, Waterboer T, Hippchen T, et al. Persistent symptoms in adult patients one year after COVID-19: a prospective cohort study. Clin Infect Dis an Off Publ Infect Dis Soc Am. 2021;.

18. Tipton CM, Fucile CF, Darce J, et al. Diversity, cellular origin and autoreactivity of antibody-secreting cell population expansions in acute systemic lupus erythematosus. Nat Immunol. 2015; 16(7):755–765.

19. Jenks SA, Cashman KS, Zumaquero E, et al. Distinct Effector B Cells Induced by Unregulated Toll-like Receptor 7 Contribute to Pathogenic Responses in Systemic Lupus Erythematosus. Immunity. 2018; 49(4):725-739.e6.

20. Ong SWX, Fong S-W, Young BE, et al. Persistent Symptoms and Association With Inflammatory Cytokine Signatures in Recovered Coronavirus Disease 2019 Patients. Open forum Infect Dis. 2021; 8(6):ofab156.

21. Phetsouphanh C, Darley D, Howe A, et al. Immunological dysfunction persists for 8 months following initial mild-moderate SARS-CoV-2 infection. medRxiv. 2021; :2021.06.01.21257759.

22. Chen Y, Wang J, Liu C, et al. IP-10 and MCP-1 as biomarkers associated with disease severity of COVID-19. Mol Med. 2020; 26(1):97.

23. Khandaker GM, Pearson RM, Zammit S, Lewis G, Jones PB. Association of serum interleukin 6 and C-reactive protein in childhood with depression and psychosis in young adult life: a population-based longitudinal study. JAMA psychiatry. 2014; 71(10):1121–1128.

24. Wium-Andersen MK, Ørsted DD, Nielsen SF, Nordestgaard BG. Elevated C-reactive protein levels, psychological distress, and depression in 73, 131 individuals. JAMA psychiatry. United States; 2013; 70(2):176–184.

25. Ortelli P, Ferrazzoli D, Sebastianelli L, et al. Neuropsychological and neurophysiological correlates of fatigue in post-acute patients with neurological manifestations of COVID-19: Insights into a challenging symptom. J Neurol Sci. 2021; 420:117271.

26. Koralnik IJ, Tyler KL. COVID-19: A Global Threat to the Nervous System. Ann Neurol. 2020; 88(1):1–11.

27. Kennedy RH, Silver R. Neuroimmune Signaling: Cytokines and the CNS. In: Pfaff DW, Volkow ND, editors. Neurosci 21st Century [Internet]. New York, NY: Springer New York; 2016. p. 1–41. Available from: https://doi.org/10.1007/978-1-4614-6434-1_174-1

28. Lorkiewicz P, Waszkiewicz N. Biomarkers of Post-COVID Depression. J Clin Med. 2021; 10(18).

29. Vlachoyiannopoulos PG, Magira E, Alexopoulos H, et al. Autoantibodies related to systemic autoimmune rheumatic diseases in severely ill patients with COVID-19. Ann. Rheum. Dis. England; 2020. p. 1661–1663.

30. Diao B, Wang C, Tan Y, et al. Reduction and Functional Exhaustion of T Cells in Patients With Coronavirus Disease 2019 (COVID-19). Front Immunol. 2020; 11:827.

31. Townsend L, Dyer AH, Naughton A, et al. Longitudinal Analysis of COVID-19 Patients Shows Age-Associated T Cell Changes Independent of Ongoing Ill-Health. Front Immunol. 2021; 12:676932.

32. Varghese J, Sandmann S, Ochs K, et al. Persistent symptoms and lab abnormalities in patients who recovered from COVID-19. Sci Rep. 2021; 11(1):12775.

33. Jabeen R, Kaplan MH. The symphony of the ninth: the development and function of Th9 cells. Curr Opin Immunol. 2012; 24(3):303–307.

34. Orologas-Stavrou N, Politou M, Rousakis P, et al. Peripheral Blood Immune Profiling of Convalescent Plasma Donors Reveals Alterations in Specific Immune Subpopulations Even at 2 Months Post SARS-CoV-2 Infection. Viruses. 2020; 13(1).

