## Supplementary Material 1 for "Persistent Autoimmune Activation and Proinflammatory State in Post-COVID Syndrome"

This appendix has been provided by the authors to give readers additional information about their work.

**Most Recent Update:** November 11, 2021.

**Persistent Autoimmune Activation and Proinflammatory State in Post-COVID Syndrome.**

**Contents**

**Supplementary Tables...…………………………………………………………………………3**

Table S1: Cellular markers used for immunophenotyping of lymphocytes by flow cytometry.............................................................................................................................3

**Supplementary Tables**

**Supplementary Table 1.** Cellular markers used for immunophenotyping of lymphocytes by flow cytometry.

| **Cell Subsets** | **Markers** |
| --- | --- |
| T cells | CD3^+^ |
| CD4+ T Cells | CD3^+^/CD4^+^ |
| Naïve CD4+ T cells | CD3^+^/CD4^+^/CD197^+^/CD45RO^-^ |
| Activated CD4+ T cells | CD3^+^/CD4^+^/CD38^+^/HLA-DR^+^ |
| Effector CD4+ T cells | CD3^+^/CD4^+^/CD197^-^/CD45RO^-^ |
| Effector memory CD4+ T cells | CD3^+^/CD4^+^/CD197^-^/CD45RO^+^ |
| Central memory CD4+ T cells | CD3^+^/CD4^+^/CD197^+^/CD45RO^+^ |
| CD8+ T Cells | CD3^+^/CD8^+^ |
| Naïve CD8+ T cells | CD3^+^/CD8^+^/CD197^+^/CD45RO^-^ |
| Activated CD8+ T cells | CD3^+^/CD8^+^/CD38^+^/HLA-DR^+^ |
| Effector CD8+ T cells | CD3^+^/CD8^+^/CD197^-^/CD45RO^-^ |
| Effector memory CD8+ T cells | CD3^+^/CD8^+^/CD197^-^/CD45RO^+^ |
| Central memory CD8+ T cells | CD3^+^/CD8^+^/CD197^+^/CD45RO^+^ |
| CD4+CD8+ T Cells | CD3^+^/CD4^+^/CD8^+^ |
| Th1 cells | CD3^+^/CD4^+^/CD194^-^/CD183^+^/CCR10^-^/CD196^-^ |
| Th2 cells | CD3^+^/CD4^+^/CD194^+^/CD183^-^/CCR10^-^/CD196^-^ |
| Th9 cells | CD3^+^/CD4^+^/ CD194^-^/CD196^+^ |
| Th17 cells | CD3^+^/CD4^+^/CD194^+^/CD183^-^/CCR10^-^/CD196^+^ |
| Th22 cells | CD3^+^/CD4^+^/CD194^+^/CD183^-^/CCR10^+^/CD196^+^ |
| Tregs | CD3^+^/CD4^+^/CD25^+^/CD127^low^ |
| B cells | CD19^+^/CD20^+/-^ |
| CD19+ CD20- B cells | CD19^+^/CD20^-^ |
| CD19+ CD20+ B cells | CD19^+^/CD20^+^ |
| Naïve B cells | CD19^+^/CD27^-^/IgD^+^ |
| Memory B cells | CD19^+^/CD27^+^/IgD^-^ |
| Classical memory B cells | CD19^+^/CD27^+^/IgD^-^/IgM^-^/CD38^+^ |
| Non-classical memory B cells | CD19^+^/CD27^+^/IgD^+^ |
| Plasmablasts | CD19^+^/CD20^-^/CD27^+^/IgD^-^/CD24^-^/CD38^+^ |
| Transitional B cells | CD19^+^/CD24^High^/CD38^High^ |

Abbreviations: CD: Cluster of differentiation; Ig: Immunoglobulin; HLA: Human leukocyte antigens; Th: T helper; Tregs: Regulatory T cells.
